# A prospective evaluation of breast thermography enhanced by a novel machine learning technique for screening breast abnormalities in a general population of women presenting to a secondary care hospital

**DOI:** 10.1101/2022.11.27.22282737

**Authors:** Richa Bansal, Sathiakar Collison, Bharat Aggarwal

## Abstract

**Objective:** Artificial intelligence-enhanced breast thermography is being evaluated as an ancillary modality in the evaluation of breast disease. The objective of this study was to evaluate the clinical performance of Thermalytix, a CE-marked, AI-based thermal imaging test, with respect to conventional mammography.

**Methods:** A prospective, comparative study performed between 15 December 2018 and 06 January 2020 evaluated the performance of Thermalytix in 459 women with both dense and nondense breast tissue. Both symptomatic and asymptomatic women, aged 30 to 80 years, presenting to the hospital underwent Thermalytix followed by 2-D mammography and appropriate confirmatory investigations to confirm malignancy. The radiologist interpreting the mammograms and the technician using the Thermalytix tool were blinded to the others’ findings. The statistical analysis was performed by a third party.

**Results:** A total of 687 women were recruited, of whom 459 fulfilled the inclusion criteria. Twenty-one malignancies were detected (21/459, 4.6%). The overall sensitivity of Thermalytix was 95.24% (95% CI, 76.18-99.88), and the specificity was 88.58% (95% CI, 85.23-91.41). In women with dense breasts (n=168, 36.6%), the sensitivity was 100% (95% CI, 69.15-100), and the specificity was 81.65% (95% CI, 74.72-87.35). Among these 168 women, 37 women (22%) were reported as BI-RADS 0 on mammography; in this subset, the sensitivity of Thermalytix was 100% (95% CI, 69.15-100), and the specificity was 77.22% (95% CI, 69.88-83.50).

**Conclusion:** Thermalytix showed acceptable sensitivity and specificity with respect to mammography in the overall patient population. Thermalytix outperformed mammography in women with dense breasts and those reported as BI-RADS 0.

**Key points:**

1. Compared with mammography as the reference, Thermalytix showed acceptable sensitivity and specificity of 95.24% and 88.58%, respectively, in the overall patient population (N=459).
2. Among women with dense breasts (n=168), 37 women (22%) were reported as BI-RADS 0 (incomplete examination) on mammograms, whereas Thermalytix identified all malignancies.
3. Thermalytix identified all malignancies among premenopausal women and women younger than 50 years of age.

**Summary Statement:** Thermalytix, an AI-based thermal imaging tool, showed acceptable sensitivity and specificity when mammography was used as the reference in the overall patient population and in women with dense breast tissue.

## INTRODUCTION

Breast cancer is the most common cancer in women worldwide, with approximately 2.26 million new cases and 684 thousand deaths reported in 2020 [1]. In developing countries such as India, there are approximately 163,000 new cases every year, with incidence and mortality rates of 25.8 and 12.7 per 100,000, respectively [2]. The problem is complicated by a critical shortage of radiologists—1 radiologist per 100,000 persons in India—indicating lack of access to expertise and widespread cancer screening facilities. This trend is seen around the globe, with fears of an impending epidemic of breast cancer mortality [3] that calls for the development of techniques other than mammography [4] to reverse this trend.

Clinical breast examination has emerged as an alternative technique in developing countries [5], although the success of its use for early screening has not been widely established. [6] Breast thermography is another adjunct screening technique that interprets heat patterns on breasts [7]. Previous usages had lower resolution due to the use of earlier generations of thermal cameras.Furthermore, the analysis of breast thermal images is complicated because asymmetric breast heat patterns are also seen with benign breast conditions. In the past, thermal images were represented using false color palettes, which required the interpreter to identify the malignancy visually from these false color images. Hence, the thermal interpretation results were highly subjective with low sensitivity and specificity. [7]

Modern high-resolution thermal cameras can detect temperature differences of 0.025 °C, and when combined with computer algorithms for thermal analysis, they may reduce subjectivity and enable automated quantitative interpretation, thereby making the interpretation process more factual [8]. Scores are generated using machine learning algorithms over medically interpretable parameters that describe the metabolic activity inside the breast tissue and indicate the presence of a possible malignancy. This mirrors the trend in the field of digital mammography, wherein the use of machine learning algorithms for extracting, detecting, characterizing and classifying radiomics features of mammograms has shown clinical benefit and are extensively used [9]. AI-enhanced breast thermography uses similar principles and is currently being re-evaluated at various centers as an ancillary modality in the screening and diagnosis of breast disease [10–13].

The objective of this prospective study is to evaluate the clinical performance of Thermalytix, a CE-marked AI-based thermal imaging test that uses machine learning on breast thermal images to generate a quantitative interpretation report, and compare it with that of conventional mammography.

## MATERIAL AND METHODS

This prospective, cross-sectional study (NCT04688086, dated 15/11/2018) was carried out at Max Super Speciality Hospital, Saket, New Delhi between 15 December 2018 and 06 January 2020 after obtaining approval from the Institutional Ethics Committee. The entire study was performed in accordance with relevant regulations, in accordance with the Declaration of Helsinki, and written informed consent was obtained from all participants.

Women who came in for breast mammography between the ages of 30 and 80 years were included in the study. Exclusion criteria were the following: pregnancy, current lactation, and previous history of breast cancer, previous lumpectomy or any active illnesses. Relevant data were obtained using a standardized pro forma.

All women included in the study underwent a Thermalytix scan followed by mammography to avoid any possible thermal disturbances created due to breast compression during mammography. Thermalytix was performed by a minimally trained technician. The mammogram was performed by a mammography technician and was reported by the senior radiologist. The results of both tests were determined independently, and each technician was blinded to the other. The resulting data were analyzed by a third-party statistician.

### Thermalytix technique

The scan was performed as per guidelines of the American Academy of Thermography [14]. A trained technician captured five thermal images from the neck to the abdomen region, namely, frontal, left-oblique, left-lateral, right-oblique, and right-lateral views. These images were then uploaded to the Thermalytix software.

The technological basis of the Thermalytix computer-aided detection engine has been published elsewhere [15–18]. Briefly, Thermalytix software automatically analyzes the uploaded images to detect abnormal patterns based on asymmetry in the structural, vascular, and thermal properties of the observed abnormality. To classify heat patterns, 31 features are extracted from abnormal regions, including boundary and shape features of hotspots and warm spots, relative temperature, symmetry, thermal distribution ratio and area differences. For vascular analysis, we convolve the thermal image with three different Gaussian functions to enhance the vessel boundaries and use shape and temperature filters to identify the pixels that form structures. Once the vessels are segmented, 21 features are extracted, including tortuosity, number of vessels, number of branches, extent of vessels, and symmetry of vessels in both breasts.

Three random forest (RF) classifiers configured for 200 decision trees over independent sets of vascular, thermal and areolar features were used to obtain the three Thermalytix scores, namely, the vascular, thermobiological and areolar scores. [19]. These scores were then combined to obtain the fourth ensemble score. ‘0’ denoted negative for malignancy, ‘1’ denoted a high likelihood of malignancy, and ‘0·5’ was used as a threshold for differentiating between negative and positive for malignancy. The logical rules used to obtain the final classification are described in Table 1.

**Table 1:** Logical rules to obtain the final classification of women positive on Thermalytix

| Age Groups | Definition of Thermalytix Positive |
| --- | --- |
| Women aged below 65 years | Thermo-biological score $\geq 0.5$,<br>Areolar score $\geq 0.5$,<br>Ensemble score $\geq 0.6$,<br>Vascular score $\geq 0.5$ and presence of lump, Ensemble score $\geq 0.48$ and lump |
| Women aged 65 and above | Thermo-biological score $\geq 0.35$,<br>Areolar score $\geq 0.35$,<br>Ensemble score $\geq 0.5$,<br>Vascular score $\geq 0.5$ and presence of lump, Ensemble score $\geq 0.48$ and lump |

Thermalytix software version 3, dated 5 December 2018, was used for the analysis.

### Mammography technique

As per the protocol, participants who underwent the Thermalytix test underwent four-view diagnostic mammography. The assessment of breast density and interpretation of images was performed by trained radiologists as per the American College of Radiology (ACR) Breast Imaging Reporting and Data System (BI-RADS). Breast density categories ‘a’ and ‘b’ were considered nondense breasts, and categories ‘c’ and ‘d’ were considered dense breasts. An assessment of BI-RADS 0, 4, and 5 was considered test-positive for mammography. The reports of the Thermalytix text were blinded to the radiologist until the mammography interpretation was completed. The Principal Investigator assessed the results to identify participants requiring additional diagnostic tests. Participants interpreted as test-positive on mammography were referred for biopsy. Where mammography was negative and the Thermalytix result was positive or when the mammographic assessment was incomplete (BI-RADS 0), breast ultrasonography (US) or MRI was recommended for further evaluation. These participants were given appointments for another day as was the standard practice at the participating hospital. All suspected disease-positive cases of malignancies were confirmed by histopathological diagnosis.

The participating clinical institution used the GE DMR Plus Analog Mammography classic CR Carestream® machine from Dec 2018 and replaced it in August 2019 with a Fujifilm AMULET Innovality. The first 330 women underwent mammography with the GE machine, and the next 129 women were studied using the Fujifilm machine.

### Statistical Analysis

The data were analyzed by Statiza Statistical Services, India. The sample size was calculated with a two-sided 95% confidence interval (CI), assuming a sensitivity of 80% and target width of 0.5 using the Clopper-Pearson Interval (Exact) Method. Considering a breast cancer prevalence of 4% in the tertiary-care center (institutional data, unpublished), approximately 450 women were estimated to be recruited. The sensitivity, specificity, positive predictive value (PPV), and negative predictive value (NPV) of the Thermalytix test in identifying breast malignancy were calculated. All statistical analyses were performed using R Software version 3·5·0 and SAS® Version 9·4 or higher [SAS Institute Inc., USA].

The study was funded by Niramai Health Analytix, developer of the Thermalytix system. However, the evaluation was done without any interference from the company. The company was not responsible for data collection, nor did it have access to or control of the data or its analysis prior to manuscript preparation.

## RESULTS

### Study population characteristics

Of the 687 women invited for screening based on the eligibility criteria, 49 women (7.1%) were excluded from analysis due to insufficient cooling during thermal imaging (based on computed temperature range), 4 (0.6%) due to a missing case files, lack of informed consent, an incomplete case report, or previous use of chemotherapy (n= 1 each), and 11 did not under go either mammography or thermography and thus did not comply with the protocol. In addition, 64 women (9.3%) had an incomplete mammographic examination (BI-RADS 0) but did not report for the recommended additional ultrasonography investigation and hence were excluded from the analysis. Another 100 participants (14.5%) who were recommended a breast ultrasonography investigation, as there was no consensus between mammography and Thermalytix reports, but still did report for the additional investigation were also excluded from the analysis. A flow diagram of the study and the test results is included in Figure 1.

**Figure 1:**
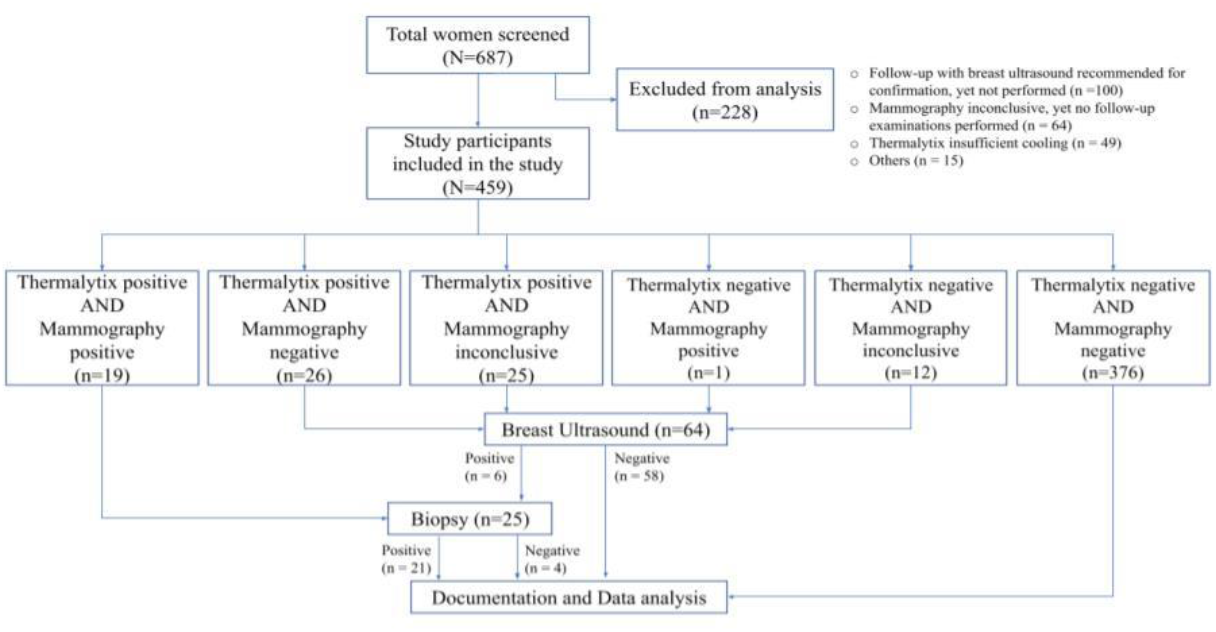
Flow diagram of the study and test results

A total of 459 women who underwent both Thermalytix scanning and mammography were included in the study analysis. The demographic characteristics were as follows. The mean age of the screened population was 50.76 (±7.3) years, with 265 (57.7%) participants being postmenopausal. In the study cohort, 168 (36.6%) women were assessed to have dense breast tissue (ACR category C–heterogeneously dense breast or category D–extremely dense breast). Sixty-nine women (15%) presented with symptoms such as breast lumps, breast pain, nipple discharge, or a combination of these symptoms.

There were 21 cases of pathology-proven breast malignancies, 19 of which were symptomatic. While 20 cases were reported as invasive ductal carcinoma, one case was reported as an intracystic papillary neoplasm with ductal carcinoma in situ.

### Overall performance of the Thermalytix system

To determine the sensitivity and specificity of the Thermalytix system, a threshold of ‘0·5’ was used, i.e., women with a score < 0.5 were categorized as negative, and those with scores between 0.5 and 1 were categorized as positive for malignancy. The Thermalytix system demonstrated an overall sensitivity of 95.24% (95% CI, 76.18-99.88), specificity of 88.58% (95% CI, 85.23-91.41), PPV of 28.57% (95% CI, 18.4-40.62), and NPV of 99.74% (95% CI, 98.58-99.99) (Figure 2 and Table 2).

**Table 2.**
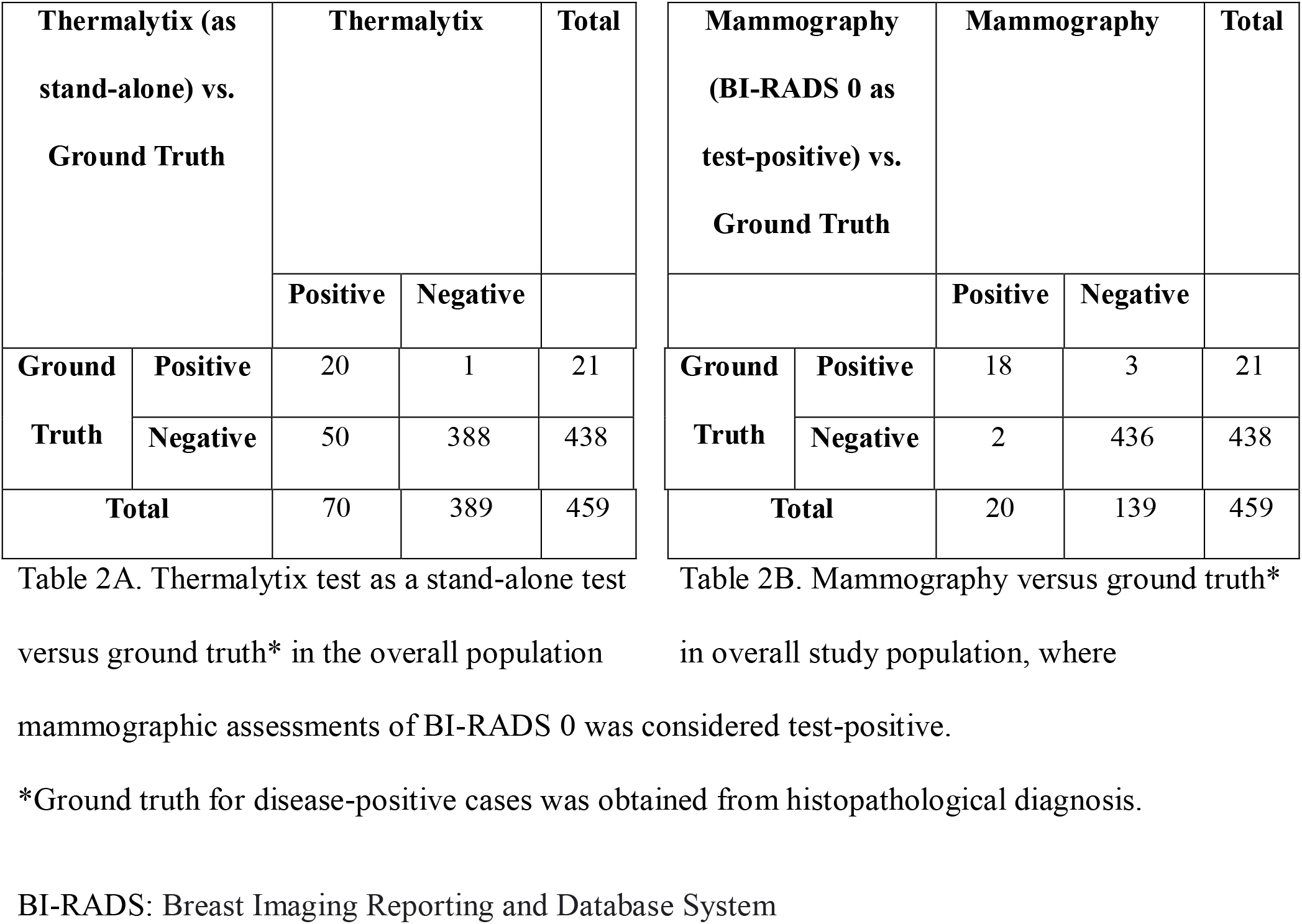
Contingency tables for overall population (N=459)

**Figure 2:**
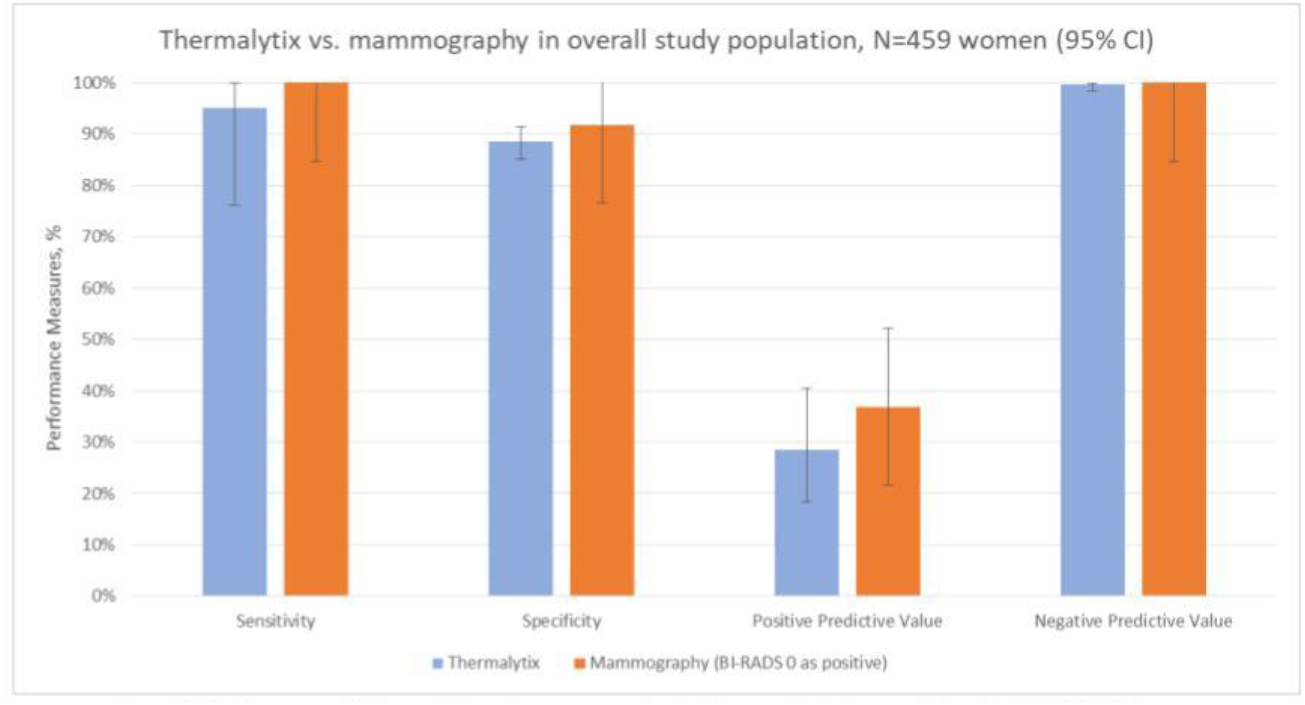
Performance of Thermalytix and Mammography in the overall study population, N=459 (95% Cl) Confidence Interval; Bl-RADS, Breast Imaging-Reporting and Data System.

Of the 21 malignancies, Thermalytix identified 20 women as positive and one woman as negative for malignancy. A receiver operating characteristic (ROC) curve for the Thermalytix system was drawn to observe the performance for different threshold points, and the area under the curve (AUC) was found to be 0·9423.

Thermalytix estimated 70 participants as test positive (70/459, 15.2%), of whom 51 women (51/70, 73%) had at least one breast abnormality on mammography or ultrasonography, warranting further investigation (Figure 3). These positive Thermalytix tests correlated with the observation of significant mammographic findings (BI-RADS 0 or 4 or 5) in a majority of patients (44 patients, 63%). Hence, the PPV of the Thermalytix for radiological positivity was 72.9% (95% CI, 62.43-83.27).

**Figure 3:**
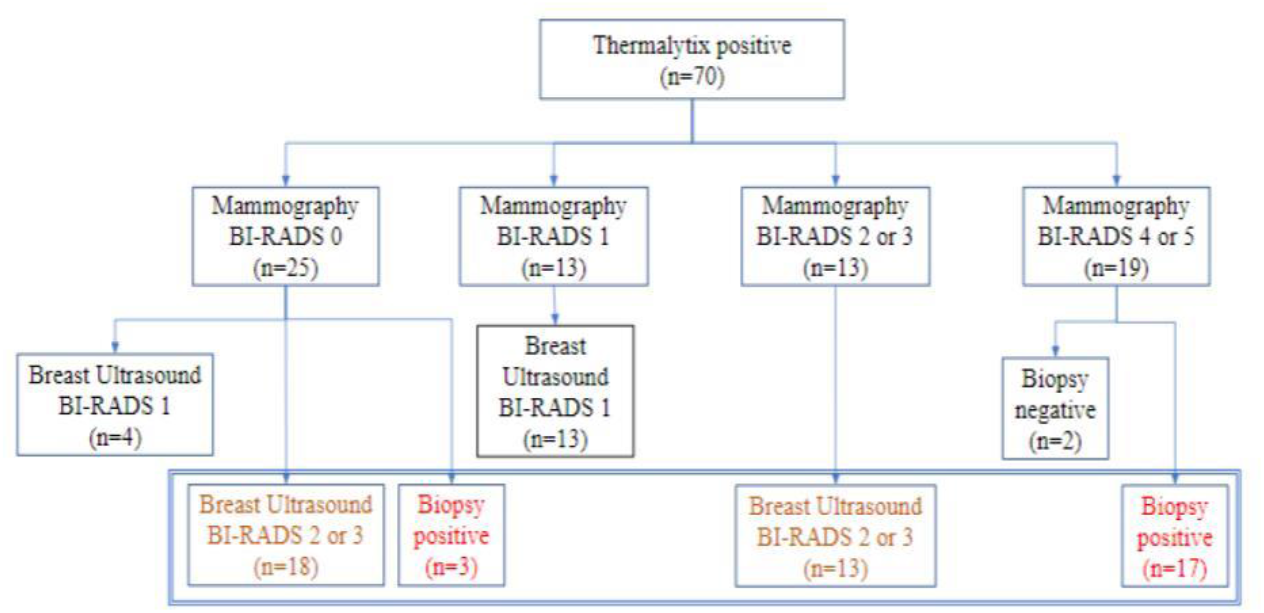
A summary of the cases identified by Thermalytix as positive.

### Performance of the Thermalytix in women with BI-RADS 0 mammographic assessments

Of the 459 women included in the study, mammography assessment was incomplete (BI-RADS 0) for 37 women (8%). After diagnostic work-up for these women with breast ultrasonography, 10 women were categorized as BI-RADS 1, 17 women as BI-RADS 2, six women as BI-RADS 3 and four women were recommended biopsies. Of the four women who underwent biopsy, three were found to be positive for malignancy. These three were also identified as positive for malignancy by the Thermalytix. When the final classification was low risk (BI-RADS 1 or 2, n=27), the Thermalytix was in agreement in 44% of patients, n=12. When the final classification was high risk (BI-RADS 3 or 4, or requiring biopsy, n=10), the Thermalytix reported high risk in all patients.

Thus, the use of Thermalytix on the 37 inconclusive examinations of mammography would have potentially eliminated the need for additional imaging investigations in 12 disease-negative cases. Considering the 37 BI-RADS 0 cases as positive, the overall sensitivity and specificity of mammography were 100% (95% CI, 83.89-100) and 91.78% (95% CI, 88.80-94.18), respectively.

### Performance in women across breast densities

In the study population, 168 (36.6%) women had dense breasts with ACR categories ‘c’ or ‘d’, which included 57% of women under the age of 45 years. In this group, the Thermalytix system had a sensitivity of 100% (95% CI, 69.15-100), specificity of 81.65% (95% CI, 74.72-87.35) (Figure 4 and Table 3), and AUC of 0.9316 (Figure 5).

**Table 3.**
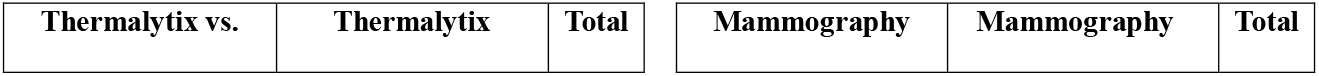
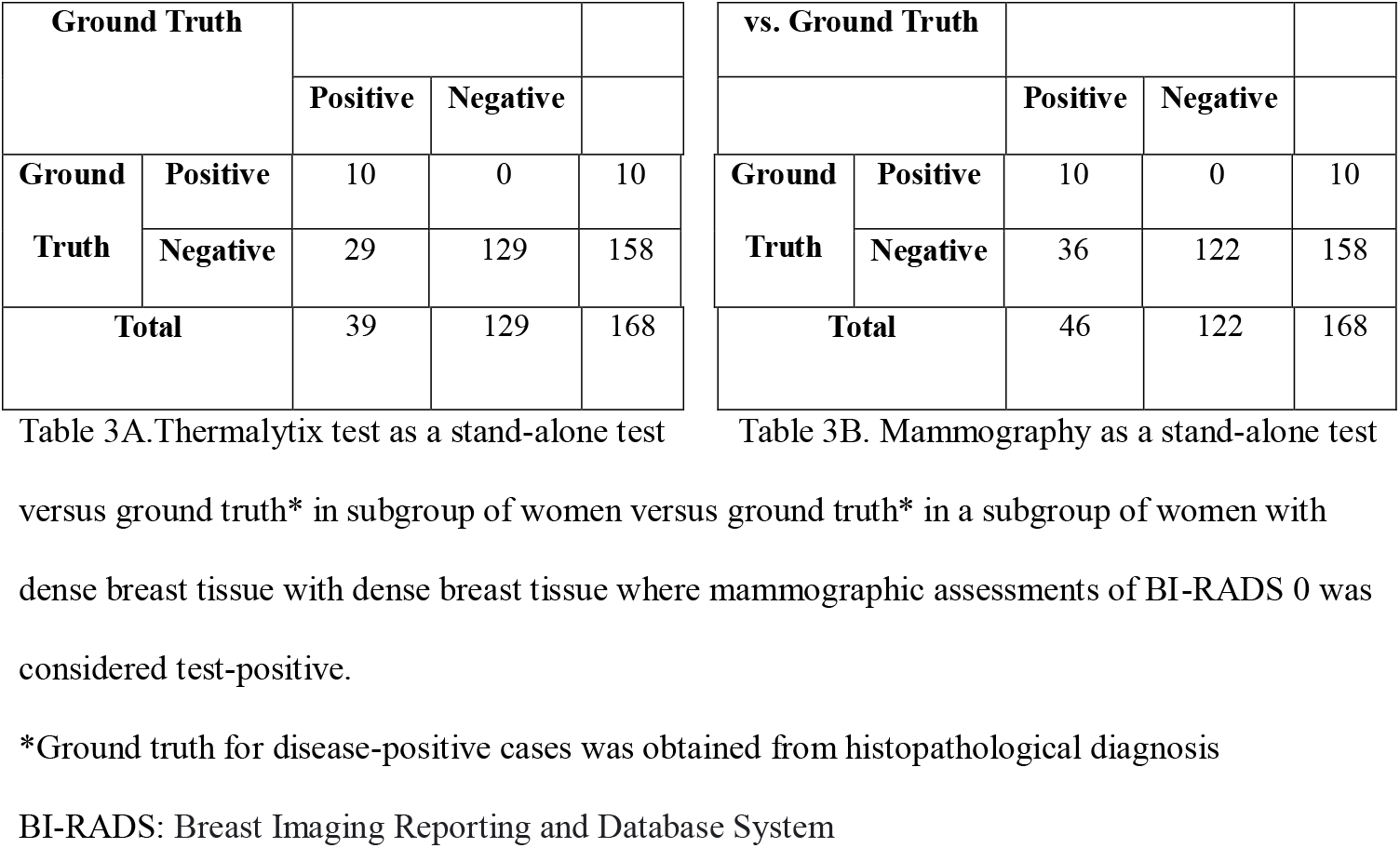
Contingency tables for women with dense breast tissue (n=168)

**Figure 4:**
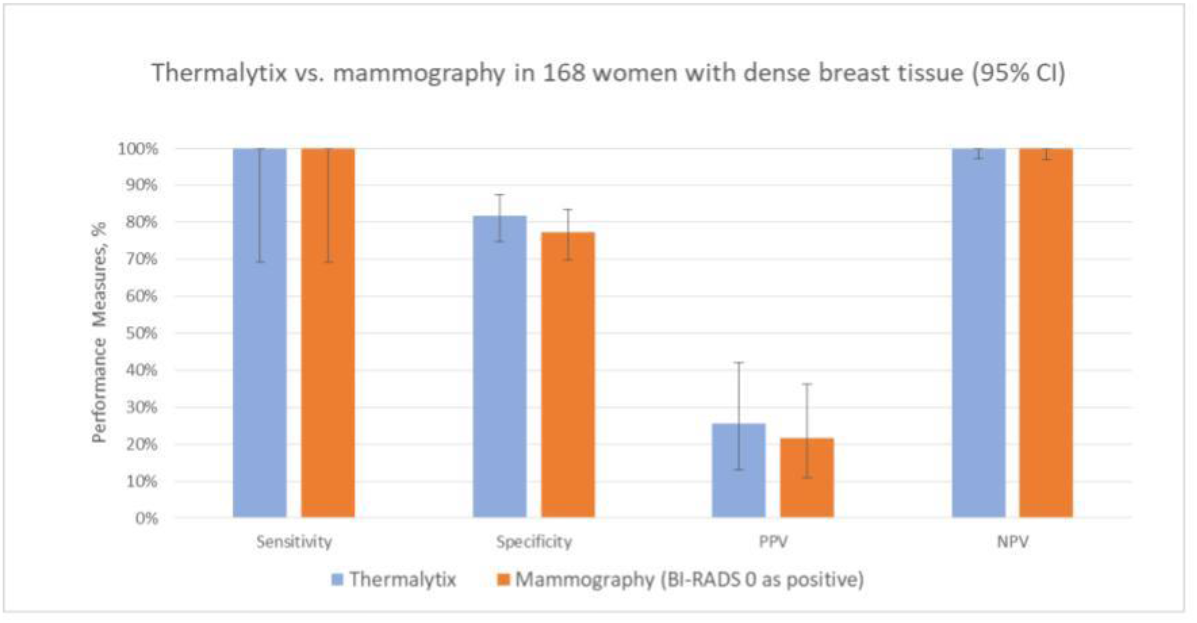
Performance of Thermalytix and Mammography in women with dense breast tissue (ACR categories ‘c’ and ‘d’), N=168 (95 % CI)

**Figure 5:**
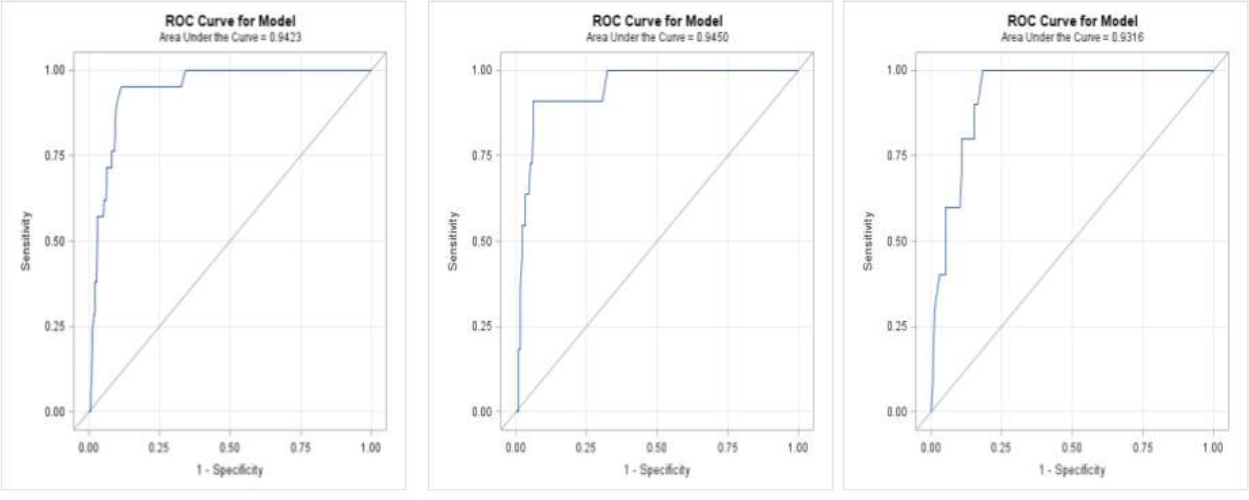
5A-Overall ROC curve for Thermalytix, N=459; 5B-ROC curve for dense breast categories ‘a’ and ‘b’, N= 291; 5C-ROC curve for dense breast categories ‘c’ and ‘d’, N=168

**Figure 6:**
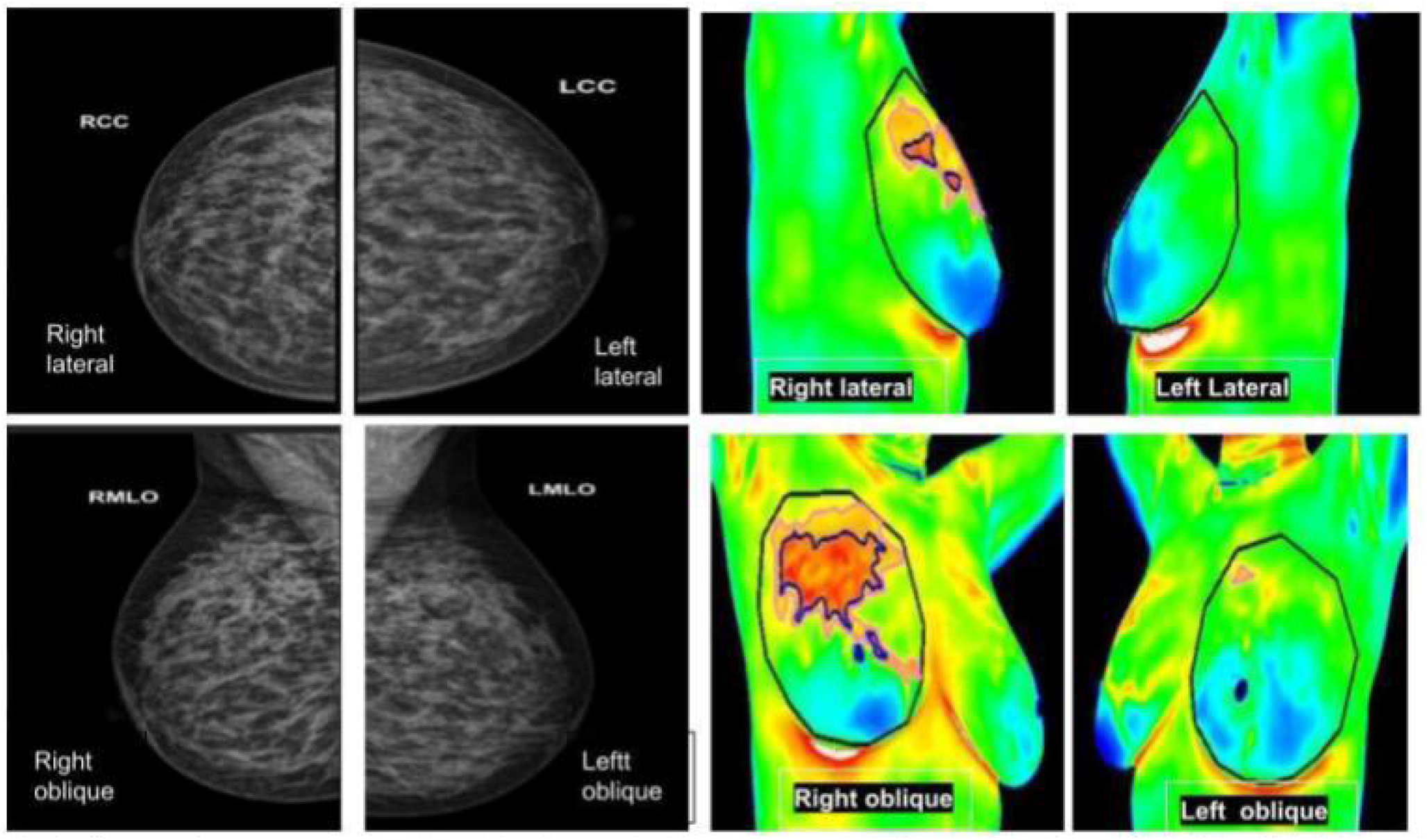
Left panel, mammography; right panel, thermography. An asymptomatic lady with dense breasts reported as BIRADS 0, in which Thermalytix revealed a suspected lesion in the upper/ outward quadrant of the right breast. Subsequently, histology was invasive ductal carcinoma

**Figure 7:**
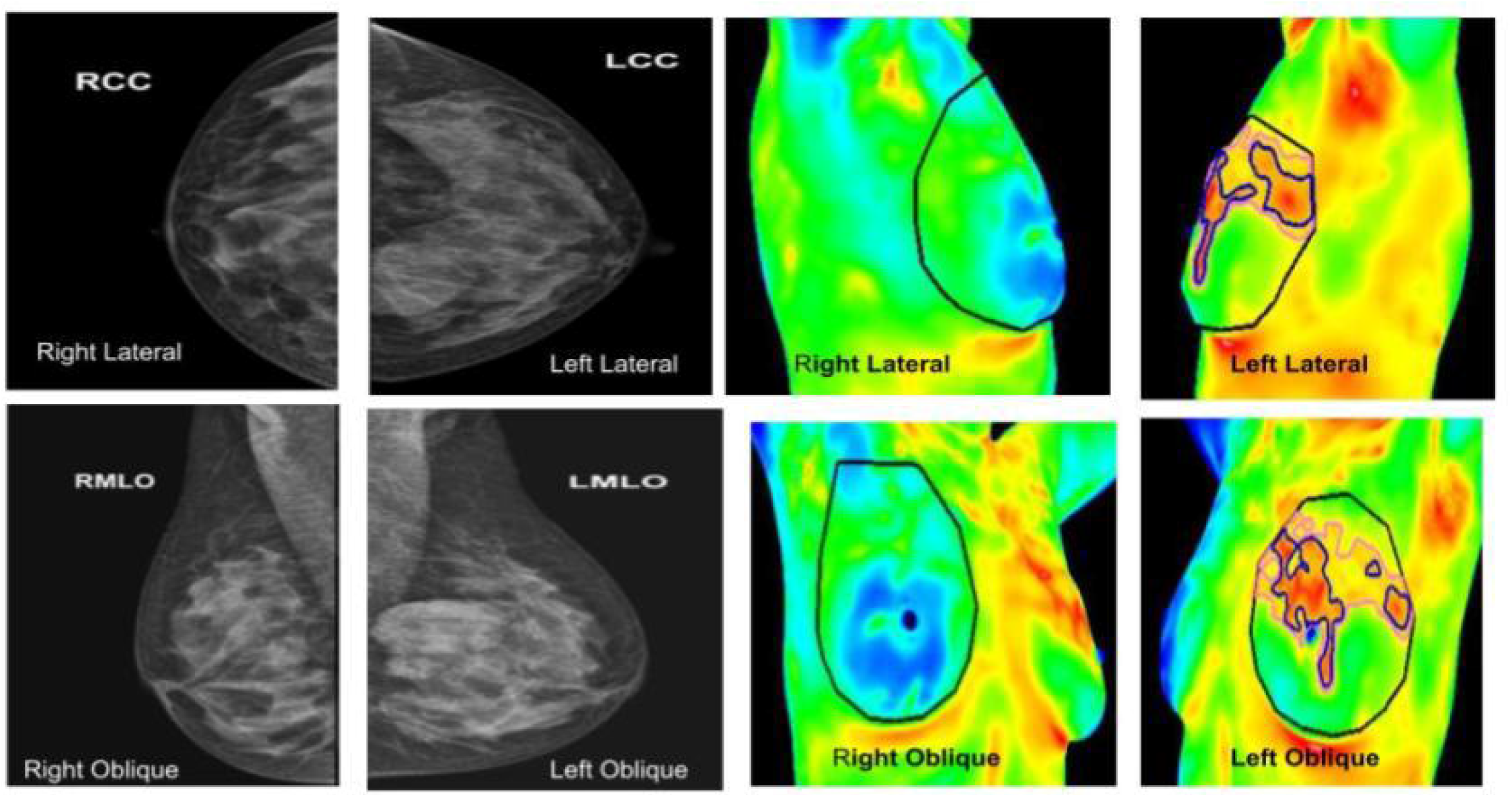
Left panel, mammography; right panel, thermography. An asymptomatic lady with dense breasts, with ill-defined opacity in the left breast on mammography, in which Thermalytix revealed a suspect lesion in the peri-areolar/ inner quadrant of right breast. Subsequently, histology was invasive ductal carcinoma

Among women with nondense or fatty breasts (n=291), the Thermalytix system had a sensitivity of 90.9% (95% CI, 73.9-100), specificity of 92.5% (95% CI, 89.4-95.6), and AUC of 0.9450 (Figure In comparison, the sensitivity and specificity of mammography in women with dense breast tissue were 100% (95% CI, 69.15-100) and 77.22% (95% CI, 69.88 to 83.5), respectively.

## DISCUSSION

The Thermalytix system demonstrated an overall sensitivity of 95.24% and specificity of 88.58% in this study of 459 participants. These values compare favorably with performance benchmarks for mammography across the world, which range from 86.9% sensitivity and 88.9% specificity in the USA [20] to 79.0% sensitivity and 96.2% specificity in Spain and 75.5% sensitivity and 97.1% specificity in Norway [21].

To the best of our knowledge, the CE-marked Thermalytix is the only technique based on thermal imaging that has prospectively studied patients to assess the role of the technology in robust clinical studies. In a previous multisite observational study of 470 symptomatic and asymptomatic women [22], the Thermalytix obtained a sensitivity of 91.02% and specificity of 82.39% with an overall area under the curve (AUC) of 0.90. In another publication [23] on a prospective multicenter study of 258 symptomatic women, an earlier version of the Thermalytix had a sensitivity of 82.5% and specificity of 80.5% with respect to the diagnostic mammogram, which had a sensitivity of 92% and specificity of 45.9%. While all the other studies of Thermalytix systems included patients who presented for either mammography or ultrasonography, in the current study, mammography was the reference standard for all participants.

In contrast, many researchers have retrospectively studied AI-based breast thermography using the publicly available Mastology Research Database, which contains thermographic images and clinical data obtained from patients of the Hospital Universitário Antônio Pedro of the Fluminense Federal University, Brazil [24]. Ramon et al. [10] used this database and tested 454 cases using their automatic model. The method showed a sensitivity and specificity of 0.8684 and 0.8943, respectively. Ruiz et al. [11] also used this database, obtaining accurate results between 90.17% and 98.33%, which are competitive with those of related works. Cauce et al. [12] applied their model to the database and achieved an AUC of 0.99, with a specificity of 100% and a sensitivity of 83%.

Sharma et al. [13] obtained a sensitivity, specificity, and AUC of 95%, 93.33%, and 95.11%, respectively, when they used a two-level hybrid method and machine learning algorithm on the database. Tayel et al. [25] used advanced convolutional networks to assess the database using 285 images and observed a sensitivity and specificity of 96.4%, 97.5%, and 97.8%, respectively. Aza et al. [26] evaluated the dataset using AlexNet in combination with support vector machines and achieved 95.56% sensitivity and 89.80% precision.

Using a different retrospective database, Kazerouni et al. [27] used a support vector machine with an RBF kernel for image retrieval and used their MATLAB model on 400 thermographic images captured and collected by Hakim Sabzevari Medical Imaging Group in Iran. The sensitivity and specificity of the model were 100% and 98%, respectively. Acharya et al. [28] used 50 infrared (IR) breast images collected from Singapore General Hospital, Singapore. Their proposed system gave an accuracy of 88.10% and sensitivity and specificity of 85.71% and 90.48%, respectively.

Gonsalves et al. [29] used a unique database of 70 images to evaluate their support vector machine-based model and obtained a specificity of 83.33% and a sensitivity of 75%.

Compliance to comprehensive breast screening programmes even in the most developed nations is far from particular due to a host of patient, economic and system-level barriers that impact screening rates especially among disadvantaged populations, such as lack of awareness, no knowledge about breast cancer, fear of the results of tests, scare of undergoing mammography, stigma of getting cancer, financial pressure, lack of time/ privacy, accessibility to treatment facilities and presence of male health workers.

Thermalytix removes some of these barriers. It is a non-contact, non-invasive, no breast compression test which is privacy aware, uses no radiation, is affordable, portable and light small screening device. As a portable device it improves access to care. It is affordable and hence, is available for all socio-economic groups. The test can be made available as the remotest of health centres, bridging geographical distances and can be conducted by low-skilled healthcare workers. The cost is lower than standard imaging modalities due to two main reasons-firstly the infrastructure involved in setting up the test is lower than standard imaging modalities; secondly it can be performed by a trained paramedical thus reducing the burden on specialists. Hence is associated with high levels of patient satisfaction. Women who are shy and sensitive to touch of private parts can use the test without inhibition.

While more large-scale studies need to be performed, this prospective study shows that the Thermalytix is emerging as a promising modality for screening women for breast cancer.In resource-constrained settings such as LMICs and developing countries, due to the absence of population-based mammography screening programs, low-cost approaches such as CBE are employed for community-based breast cancer screening programs [30]. However, CBE can detect only palpable lesions and has a low sensitivity of 28% to 54% [6]. In comparison, the AI-based Thermalytix test is automated, nonsubjective and has demonstrated a higher sensitivity of 95.24% and thus could prove to be an affordable method for downstaging the disease at presentation.We acknowledge the limitations of the current study. There were many participants who were lost to follow-up. Out of the 687 women recruited for the study, 228 women were excluded for the following reasons: 107 women had a BI-RADS 0 assessment on mammography, and only 37 of them completed the study protocol of the recommended follow-up with US. This loss to follow-up of study participants (n=70) with incomplete mammographic assessments is a common occurrence and is usually addressed by double reading of mammograms to obtain conclusive observations in many countries. Since double reading was not in our approved study protocol, we could not consider the same.

Another reason for the dropouts was erroneous thermal imaging with insufficient cooling of participants before capturing the thermal images (n=49). When a root-cause analysis was performed, two potentially correctable issues were found that will be addressed in future studies. 1) It was found that intermittent problems in the centralized air-conditioning system caused sudden temperature fluctuations in the examination room that affected the precooling of some participants and consequentially reduced the quality of captured thermal images and led to protocol nonconformance. To address this limitation, a simple portable water cooler system is now being used to ensure the appropriate temperature in the room. 2) Additionally, to help the technician in capturing good images, an innovative ‘image check’ feature was later incorporated into the Thermalytix software to automatically identify image quality issues and alert the technician of the same during live image capture and screening.

## CONCLUSION

This study compared the effectiveness of Thermalytix in detecting breast cancers in symptomatic and asymptomatic populations. While the results of the study are promising, future studies on a large population will aid in gathering more evidence and in understanding the placement of Thermalytix systems in the breast cancer care pathway.

## Data Availability

All data produced in the present study are available upon reasonable request to the authors

## Abbreviations

AI: Artificial Intelligence
ACR: American College of Radiology
AUC: Area under the curve
BI-RADS: Breast Imaging Reporting and Data System
CBE: Clinical breast examination
LMICs: Low-and middle-income countries
PPV: Positive-predictive value
NPV: Negative-predictive value
ROC: Receiver operating characteristic
RF: Random Forest

## Declaration Section

### Ethical Approval and Consent for participation

This prospective, cross-sectional study was carried out at Max Super Speciality Hospital, Saket, New Delhi after obtaining approval from the Institutional Ethics Committee, Max Super Speciality Hospital, Saket, New Delhi. The trial was registered with The Clinical Trials Registry-India, number: NCT04688086 available at the ICMR’s National Institute of Medical Statistics (http://icmr-nims.nic.in). We state that written informed consent was obtained in writing from all the study participants.

### Consent for publication

We wish to confirm that all authors have approved the manuscript for submission, and have consented for publication. Also, we wish to confirm that the content of the manuscript has not been published, or submitted for publication elsewhere

## Availability of data and materials

The datasets generated during and/or analysed during the current study are available from the corresponding author on reasonable request.

## Authors’ contributions

RB: Conceptualization, Data curation, Investigation, Methodology, Project administration, Resources, Supervision, Validation, Visualization, Writing – review & editing

BA: Conceptualization, Methodology, Resources, Supervision, Validation, Writing – review & editing

SC: Formal Analysis, Visualization, Writing – review & editing

## Acknowledgements

Not applicable

